# SARS-CoV-2 screening prevalence in educational staff in Berlin, Germany, June-December 2020

**DOI:** 10.1101/2021.05.19.21257452

**Authors:** Sophia Kindzierski, Welmoed van Loon, Stefanie Theuring, Franziska Hommes, Eberhard Thombansen, Malik Böttcher, Harald Matthes, Heike Rössig, David Weiger, Christof Wiesmann, Tobias Kurth, Valerie Kirchberger, Joachim Seybold, Frank P Mockenhaupt, Maximilian Gertler

## Abstract

SARS-CoV-2 infections in childcare and school settings potentially bear occupational risks to educational staff. We analyzed data derived from voluntary, PCR-based screening of childcare educators and teachers attending five testing sites in Berlin, Germany, between June and December 2020.

Within seven months, 17,491 tests were performed (4,458 educators, 13,033 teachers). Participants were largely female (72.9%), and median age was 41 years. Overall, SARS-CoV-2 infection prevalence was 1.2% (95%CI, 1.1-1.4%). Prevalence in educational staff largely resembled community incidence until the start of the second pandemic wave in mid-September 2020, when an unsteady prevalence plateau was reached. Then, infection prevalence in teachers (1.2% [95%CI, 0.8-1.8%]) did not significantly differ from the population prevalence (0.9% [0.6-1.4%]) but it was increased in educators (2.6% [1.6-4.0%]; aOR, 1.6 [1.3-2.0]). Irrespective of occupation, those that reported contact to a confirmed SARS-CoV-2 case outside of work had increased risk of infection (aOR, 3.0 [95%CI, 1.5-5.5]). In a step-wise backwards selection, the best set of associated factors with SARS-CoV-2 infection involved age, occupation, and calendar week.

These results are in line with findings that teachers do not bear an increased risk of SARS-CoV-2 infection, while childcare educators do. Infection control and prevention measures need to be strengthened in child care settings to further reduce respective occupational hazards. At the same time, the private environment appears to be the main source of SARS-CoV-2 infection for educational staff.

## Introduction

The operation of schools and childcare centers during the COVID-19 pandemic is subject to intense debate. Educators and teachers are front-line workers in the education system. Considerations of the children’
ss contagiousness and potential transmission chains have led to concerns about safe school attendance and workplace security. Such concerns, in turn, impact on the functionality of schooling and childcare [1]. While the influence of opened or closed educational facilities on the course of the pandemic has been subject to modelling [2] and observational studies [3–4], with partially contradicting findings, the actual occupational risk of educational staff is less clear. Repeatedly changing infection prevention and control (IPC) measures, such as to mask wearing obligations or divided classes, also make it difficult to draw firm conclusions.

The present study aimed at assessing the prevalence of SARS-CoV-2 infections in teachers and educators taking advantage of a screening offer for asymptomatic school and daycare staff in the period between June and December 2020 at five testing sites throughout Berlin. Moreover, we further aimed to evaluate the influence of occupation (teachers *vs*. educators) and recent contact to a SARS-CoV-2 case on SARS-CoV2 infection. Lastly, we aimed to identify the factors that best describe SARS-CoV2 infection risk

## Methods

### Study design, setting and participants

The Senate of Berlin initiated a free voluntary screening for asymptomatic school and daycare staff between 8 June and 31 December 2020 at five testing sites in Berlin, i.e., Charité – Universitätsmedizin Berlin (”A”; 8 June – 13 December); Vivantes Wenckebach-Klinikum (“B”; 6 July – 30 December); Vivantes Klinikum Spandau (“C”; 6 July – 30 December); Vivantes Prenzlauer Berg (“D”; 6 July – 30 December); and Gemeinschaftskrankenhaus Havelhöhe (“E”; 28 July – 30 December). School and daycare staff in Berlin, including permanent staff, trainees, and volunteers were invited to participate. The respective invitations and detailed information were sent to all schools and kindergartens and forwarded to the facilities’ staff. The screening consisted of an RT-PCR-based SARS-CoV-2 test and questionnaire-based collection of information regarding the participants’ medical situation and occupational context. Persons mildly symptomatic at presentation or primary contacts of positively tested individuals were not rejected. Repeat tests were allowed at a monthly interval. The study was reviewed and approved by the Charité ethics committee (EA1/313/20), and participants provided informed written consent.

From June to August 2020, the weekly incidence of COVID-19 cases (official notification data) in Berlin was low, i.e., around 12/100,000 inhabitants. Starting mid-September, incidence sharply increased, resulting in a peak between mid-November and mid-December 2020 with figures of up to 236/100,000 habitants (week 47) (Figure 1). With increasing community incidence, progressive IPC measures became mandatory for the population, such as facemask obligations or access limitations in certain public locations. By November 2020, a “lockdown light” was implemented comprising closure of gastronomic and cultural places as well as restrictions for social meetings and travelling. During the study period, schools and kindergartens remained continuously open for symptomless children except for school holidays. School operation, with basic hygiene measures in place, was also influenced by increasing community incidence leading to, e.g., expansion of facemask wearing from way-to-class to use-in-class. As of end of October 2020, obligatory facemask use in and outside of the classroom, split lessons, and open-window ventilation were decreed. In childcare, restrictions were less strict and included mainly regulation of intra-staff and parental contacts, but no obligatory mask wearing for children or for educators. A full lockdown was imposed on 16 December 2020, closing schools, kindergartens and cultural institutions but leaving offices and other work spaces open.

**Figure 1.**
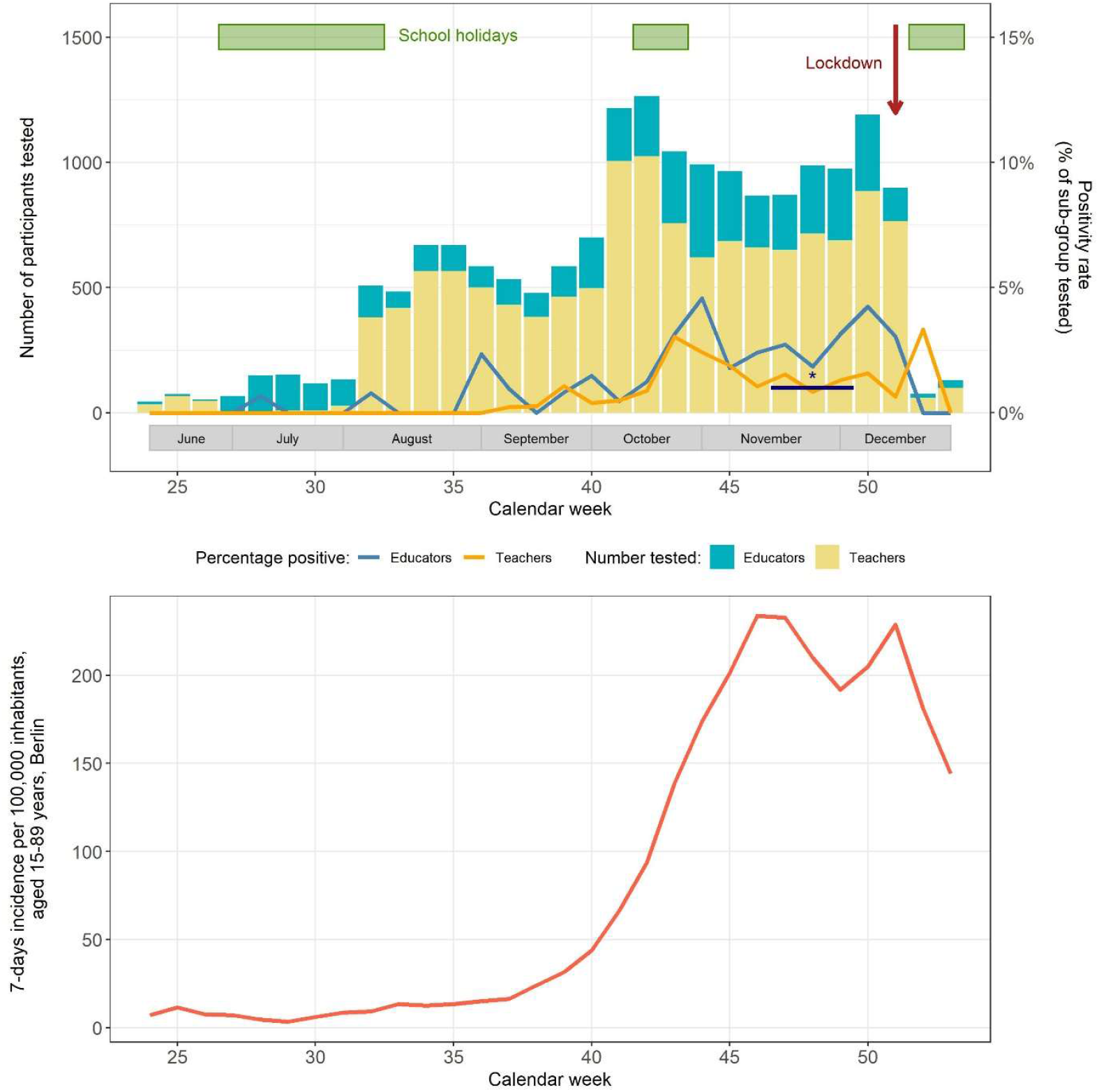
Prevalence of SARS-CoV-2 infection among educational staff and community incidence ^1^ in Berlin, June to December 2020. ^*^, community prevalence in central Berlin indicated by a line, also representing the respective sampling period.^1^ Data provided by the Senate of Health and Social Affairs, Berlin (LAGeSo) (https://www.berlin.de/corona/lagebericht/desktop/corona.html)

### Data collection and analysis

Basic demographic data (age, sex, occupation) and PCR-test results were collected. Further data were collected at two sites only, A and E, paper-based or *via* web-app, and using different questionnaires. These data included current symptoms, previous contacts to a positive case, and IPC measures.

From each participant, a nasopharyngeal, oropharyngeal or combined swab was taken by a health care professional, and SARS-CoV-2 infection was assessed by RT-PCR (cobas® 6800/8800; Roche Diagnostics, Mannheim, Germany; at site E also AltoStar®, Altona Diagnostics, Hamburg, Germany). According to German infection prevention laws, a positive test result was reported to the local health authority.

Attendees identified as working in educational settings, but not as teachers or educators, were excluded from analysis. Basic characteristics were described by their median and range, proportion, odds ratio (OR) and 95% confidence interval (CI), as applicable. We estimated the effects of occupation and recent contact to a confirmed SARS-CoV-2 case on SARS-CoV-2 infection using a binomial logistic regression model with sex, age and calendar week as covariates and random intercept effects for the five testing sites. We explored the combination of variables in the dataset that best described SARS-CoV-2 infection status with a backwards stepwise selection using the Akaike information criterion. The likelihood of variable selection was further evaluated by repeating the variable selection using bootstrap technique (Supplement 1). All analyses were done in R version 3.6.3.

## Results

Between 8 June and 31 December 2020, 18.941 tests for SARS-CoV-2 were performed. A total of 17,491 participants were included for analysis (teachers, 13,033; educators, 4,458; *per* site: A, 2,288; B, 4,553; C, 2,916; D, 4,436; E, 3,298).

At site A, where this was recorded, 15% (286/1936) of participants were repeatedly tested. Symptom data was collected at sites A and E, where 5.4% (301/5,554) of participants reported to have at least one symptom compatible with SARS-CoV-2 infection.

Participants were largely female (72.9%, 12,710/17,429), and the median age was 41 (range, 15-89) years. More educators than teachers were female (82.2% *vs*. 69.7%), and educators were younger (median age, 39 [range, 15-73] *vs*. 41 [15-89] years). Sex, age, and occupation differed only slightly between testing sites (Supplement table 1).

During the study period, 1.2% (95%CI, 1.1-1.4%; 213/17,491) of tests were positive for SARS-CoV-2, relatively evenly distributed across testing sites (1.0-1.7%;). At the two sites documenting, positively tested staff in 16.4% (11/67) reported symptoms as compared to 5.3% (290/5487) in those negatively tested.

Figure 1 displays SARS-CoV-2 prevalence in educational staff and community incidence over time. The infection prevalence in teachers (overall, 1.0% [95%CI, 0.8-1.2%], Table 1) was zero until mid-September (week 38), when it started to increase to reach a preliminary peak (3.0% [1.9-4.5%] by mid-October (week 43). In the smaller group of educators, prevalence (overall, 1.9% [1.5-2.4%], Table 1) exceeded zero at several points until mid-September, then also increased and peaked in mid-October (4.5% [2.7-7.2%]). These prevalence increases largely paralleled community incidence. Yet, in both teachers and educators, prevalence dropped 2-3 weeks before the highest community incidence, reached an unsteady four-week plateau and peaked again at the time a lockdown was implemented. During the plateau-phase (weeks 47-48), representative, unweighted adult population prevalence in the central city district was 0.9% (95%CI, 0.6-1.4%; 21/2287) [5]. Compared to that, the simultaneous prevalence in teachers was similar at 1.2% (0.8-1.8%; 24/1936,) but it was significantly increased among educators (2.6% [1.6-4.0%]; 19/741; OR, 2.8 [1.5-5.3]).

**Table 1.** Factors associated with SARS-CoV-2 infection among educational staff in Berlin, Germany, June to December 2020.

|  | Result of SARS-CoV-2 PCR |  | OR (95%CI) |
| --- | --- | --- | --- |
|  | Positive (n=227) | Negative (n=18,230) |  |
| Sex |  |  |  |
| Male | 1.3% (64) | 98.7% (4,932) | 1 |
| Female | 1.2% (162) | 98.8% (13,238) | 1.0 (0.7-1.3) |
| Age, years (median, range) | 36.0 (16.0-65.0) | 41.0 (15.0-89.0) | 0.96 (0.95-0.98) |
| Profession |  |  |  |
| Teacher | 1.0% (132) | 99.0% (13,258) | 1 |
| Educator | 1.9% (95) | 98.1% (4,972) | 2.0 (1.5-2.7) |
| Reported contacts to SARS-CoV-2 positive individuals outside work<sup>a</sup> |  |  |  |
| No | 1.4% (29) | 98.6% (2,098) | 1 |
| Yes | 6.9% (9) | 93.1% (121) | 6.4 (2.4-15.7) |
| Reported contacts to SARS-CoV-2 positive individuals at work<sup>a</sup> |  |  |  |
| No | 1.6% (29) | 98.4% (1,813) | 1 |
| Yes | 2.2% (9) | 97.8% (408) | 0.9 (0.3-2.4) |

OR, odds ratio; 95%CI, 95% confidence interval; aOR, adjusted odds ratio; <sup>a</sup> Data from one testing site (A) only, the effects of primary contacts at and outside of work on the likelihood of SARS-CoV-2 infection were estimated with a logistic regression model including age and sex as covariates.

We observed increased odds for educators, as well as for staff with reported non-work related contacts to positively tested individuals and for younger staff (Table 1). The latter was true for both educators (contacts outside of work, OR, 4.2 [95%CI, 1.4-12.4]; young age, OR, 0.97 [0.95-0.99]) and teachers (contacts outside of work, OR, 10.6 [2.7-41.7]; young age, OR, 0.97 [0.95-0.98]). When estimating the effect of occupation (teacher *vs*. educator) and contact history, we found that reported contacts to a positively tested person at work did not affect the odds of SARS-CoV-2 infection (aOR, 0.9 [95%CI, 0.4-1.8]), but contacts beyond the workplace tripled the odds (aOR, 3.0 [1.5-5.5]).Being an educator increased the odds as well (aOR, 1.6 [1.3-2.0]).

The best set of associated factors with SARS-CoV-2 infection involved age (*P*<0.001), occupation (*P*<0.001), and calendar week (*P*<0.001). The bootstrap repetition resulted in a selection frequency of each of these variables as follows: age, 99%; occupation, 99%; calendar week, 100%.

## Discussion

Our screening data spanning half a year, in which the COVID-19 pandemic developed from a low level to an intense second wave, suggest an elevated SARS-CoV-2 infection risk in daycare educators, whereas this was not obvious among teachers. This is based on the increased infection prevalence among educators (but a similar one in teachers) as compared to representative adult community prevalence [5] in November-December 2020. Notably, prevalence among educational staff increased with increasing incidence in the community, but not nearly to the same extent.

The increased SARS-CoV-2 prevalence in educators may reflect routine physical contact to children, and thus increased potential viral exposure. Concurrently, distancing or mask rules are less strictly set for educators. Beyond occupational exposure, socio-economic status and SARS-CoV-2 infection are inversely associated [6]. Becoming an educator in Germany does not require university education and usually yields a below-average salary. Moreover, educators were rather young, and age was among the set of variables associated with infection. Presumably, this goes along with more intense social activity at younger age.

Data from the UK show a SARS-CoV-2 infection incidence in teachers which is identical to working adults [7]. In the present study, infection prevalence in teachers was slightly higher than in a representative sample of the adult Berlin population [5], but confidence intervals largely overlapped. In another study at peak transmission in Berlin, early November 2020, 1.4% of 140 teachers were found to be infected [4], confirming our findings.

Infection prevalence in educational staff did not increase to the extent community incidence did in the second pandemic wave. Conceivably, this might reflect a protective impact of the educational job setting under IPC conditions, including strict quarantine implementation. An indirect proof of tolerable working conditions comes from contacts to positive persons in the private (but not the occupational) context being the factor most strongly associated with SARS-CoV-2 infection. Likewise, in autumn 2020, Austrian teachers in up to 90% were the origin of SARS-CoV-2 school clusters, contrasting their relative minority at school [8]. This underlines the necessity of social life adjustments but also of further occupational IPC measures. Infection risks stemming from the private context do not argue against the recently introduced SARS-CoV-2 testing twice a week as such will detect infections otherwise introduced into educational facilities. Increased infection prevalence in educators supports their prioritized vaccination.

Our study has several limitations including approximate proportions of 15% repeatedly tested and 5% symptomatic individuals. Under-reporting of symptoms (presumed to lead to exclusion from screening) and over-reporting (misunderstood test indication) cannot be excluded. Selection bias due to easily accessible testing might apply. At two testing sites, educational staff not being teacher or educator was excluded, and it is possible that a similar proportion of some 15% applies to the remaining data set. Nevertheless, re-running the analysis with and without exclusion (Supplement 3) did not yield differing results suggesting that our overall results are not substantially affected. Comparing prevalence to community incidence data has inherent limitations. Community incidence is based on notified symptomatic patients or primary contacts, the detection of which is subject to changing testing indication, access to testing, and test willingness. Translating the peak weekly incidence of >200/100,000 in Berlin to a prevalence over seven days of approximately 0.2% provides a figure, which is substantially lower than the simultaneous population prevalence of 1% [5]. Antibody assessments reveal incidence data to be grossly underestimated [9]. Therefore, we compared our figures with population prevalence rather than incidence. The resulting temporal restriction is deplorable as is the mere absence of representative prevalence data in Germany. Absence of complete datasets for all participants might have impacted the detection and estimates of risk factors. These limitations need to be balanced against our large sample size almost representing one fourth of the educational staff in Berlin.

## Data Availability

Corresponding author: Dr. med. Maximilian Gertler; Institute of Tropical Medicine and International Health, Charite Universitaetsmedizin Berlin, Augustenburger Platz 1, 13353 Berlin, Germany;

Analysis code is available upon request from Welmoed van Loon

## Conclusions

Educators but not teachers appeared to be at increased risk of SARS-CoV-2 infection in the second half of 2020 in Berlin. Particularly when considering the recent replacement of the original virus by the more transmissible B.1.1.7 variant in Berlin [10], occupational and private-life IPC measures need to be sustained and strengthened among educational staff while repeat-testing of staff and vaccination roll-out should be consistently pursued.

## Declarations

### Availability of data and material

The identified data is available upon reasonable request from the corresponding author.

### Code availability

Analysis code is available upon request from Welmoed van Loon

### Ethics approval

This study was performed in line with the principles of the Declaration of Helsinki. Approval was obtained from the ethics committee. The study was reviewed by the ethics committee at Charité – Universitätsmedizin Berlin, Germany (EA1/313/20).

### Consent to participate

Written informed consent was obtained from all individual participants included in the study

### Consent to publish

Written informed consent was obtained from all individual participants included in the study.

## Acknowledgements

We thank the staff at the testing sites at Charité - Universitätsmedizin Berlin, Vivantes Hospital Group, and Gemeinschaftskrankenhaus Havelhöhe. We also thank Martin Kerger and the Data4Life Team for the development of technical solutions.

