## Supplemental Material for "SARS-CoV-2 screening prevalence in educational staff in Berlin, Germany, June-December 2020"

Charité – Universitätsmedizin Berlin, corporate member of Freie Universität Berlin and Humboldt- Universität zu Berlin, Berlin, Germany; [1] Medical Directorate; [2] Institute of Tropical Medicine and International Health; [5] Institute of Public Health
[3] Vivantes Hospital Group, Berlin, Germany
[4] Gemeinschaftskrankenhaus Havelhöhe, Berlin, Germany

Corresponding author: Dr. med. Maximilian Gertler; Institute of Tropical Medicine and International Health, Charité – Universitätsmedizin Berlin, Augustenburger Platz 1, 13353 Berlin, Germany;

**Supplemental methods for backwards stepwise selection procedure**

We explored which combination of variables in our dataset best described the SARS-CoV-2 infection in a logistic regression applying backward stepwise selection by Akaike information criterion (AIC). The following variables were included for this process: SARS-CoV-2 infection status (positive/negative), sex (male/female), age (years), testing site (A/B/C/D/E) and time (by calendar week).

We fitted a full binomial logistic regression model including all variables above, with SARS-CoV-2 infection as outcome variable. Then we used this model for backwards selection by AIC (R function *stepAIC* from the *MASS* package in R). For the bootstrap analysis, this process was repeated on 1,000 resampled datasets (R function *boot.stepAIC* from the *bootStepAIC* package in R) as follows: a new dataset was simulated by resampling with replacement; the full binomial logistic regression model was fitted; the backwards stepwise selection by AIC was done on the new full model; among the 1,000 resulting selected models, the number of times was counted that each variable was selected.

All analyses were done in *R version 3.6.3* and the code is available upon request from Welmoed van Loon,.

**Supplemental Table 1**

| | Testing site | | | | |
|---|---|---|---|---|---|
| | **A N=2288 (%, [n])** | **B N=4553 (%, [n])** | **C N=2916 (%, [n])** | **D N=4436 (%, [n])** | **E N=3298 (%, [n])** |
| SARS-CoV-2 positivity | 1.3% (30) | 1% (45) | 1.7% (50) | 1.1% (51) | 1.1% (37) |
| Occupation | | | | | |
| - Teacher | 47.9% (1095) | 81.9% (3728) | 78.1% (2276) | 77.5% (3439) | 75.7% (2495) |
| - Educator | 52.1% (1193) | 18.1% (825) | 21.9% (640) | 22.5% (997) | 24.3% (803) |
| Age, median (range) | 38 (16-73) | 42 (16-73) | 40 (15-89) | 41 (16-70) | 42 (15-83) |
| Female | 74.3% (1699) | 73.8% (3357) | 70.9% (2067) | 72% (3193) | 73.8% (2394) |
| Reported symptoms | 10.4% (239) | n.a. | n.a. | n.a. | 1.9% (62) |

**Supplemental analysis**

One of the constraints in our big dataset was the lack of more detailed information on the function that staff in educational facilities fulfill for most participants. At two testing sites, educational staff not being teacher or educator could be identified and excluded. However, it is possible that a similar proportion of some 15% applies to the remaining data set. To investigate the impact of this “noise” in the occupation variable, we repeated the analyses for effect estimation of occupation and contact history on SARS-CoV-2 infection status in two data subsets: data collected at site A and E *with* and *without* exclusion of non-teacher and non-educator participants.

Excluded participants did not differ from the included participants as for SARS-CoV-2 infections status, sex, age, reported symptoms, and contact history. However, the excluded participants were more frequently categorized as educator than teacher (Supplemental Table 2) was. The outcomes of our models were almost similar with and without exclusion of non-teachers and non-educators (Supplemental Figure 1).

**Supplemental Table 2. Characteristics of excluded participants and included participants from testing site A and E. Participants were excluded because they could be identified to not be a teacher or educator, *e.g.*, facilitating staff in educational facilities.**

| | **Excluded participants N=988 (n [%])** | **Included participants N=5599 (n [%])** | **OR (95% CI)** |
|---|---|---|---|
| SARS-CoV-2 infection status | | | |
| - negative | 973 (15.0%) | 5498 (85.0%) | 1 |
| - positive | 14 (17.3%) | 67 (82.7%) | 0.9 (0.5-1.7) |
| Occupation before inspection of additional data | | | |
| - Teacher | 381 (9.6%) | 3592 (90.4%) | 1 |
| - Educator | 607 (23.2%) | 2007 (76.8%) | 0.4 (0.3-0.4) |
| Age (median, range) | 39.0 (18.0-85.0) | 41.0 (15.0-83.0) | n.a. |
| Sex | | | |
| - Male | 282 (16.3%) | 1443 (83.7%) | 1 |
| - Female | 706 (14.7%) | 4105 (85.3%) | 1.1 (1.0-1.3) |
| Any symptoms | | | |
| - no | 937 (15.1%) | 5265 (84.9%) | 1 |
| - yes | 51 (14.4%) | 302 (85.6%) | 1.1 (0.8-1.5) |
| Contact to SARS-CoV-2 case outside of work | | | |
| - no | 596 (28.0%) | 1532 (72.0%) | 1 |
| - yes | 28 (21.5%) | 102 (78.5%) | 1.4 (0.9-2.3) |
| Contact to SARS-CoV-2 case at work | | | |
| - no | 528 (28.6%) | 1315 (71.4%) | 1 |
| - yes | 96 (23.0%) | 321 (77.0%) | 1.3 (1.0-1.7) |

**Supplemental Figure 1. Comparison of effect estimates for SARS-CoV-2 infection in a dataset *with* and *without* exclusion of non-teachers and non-educators (e.g., facilitating staff in educational settings).** Effect estimates for occupation (teacher/educator) were obtained by a logistic regression including sex, age and calendar week as covariates. For the variables on contact history, the covariate set included occupation, sex, age, and calendar week.

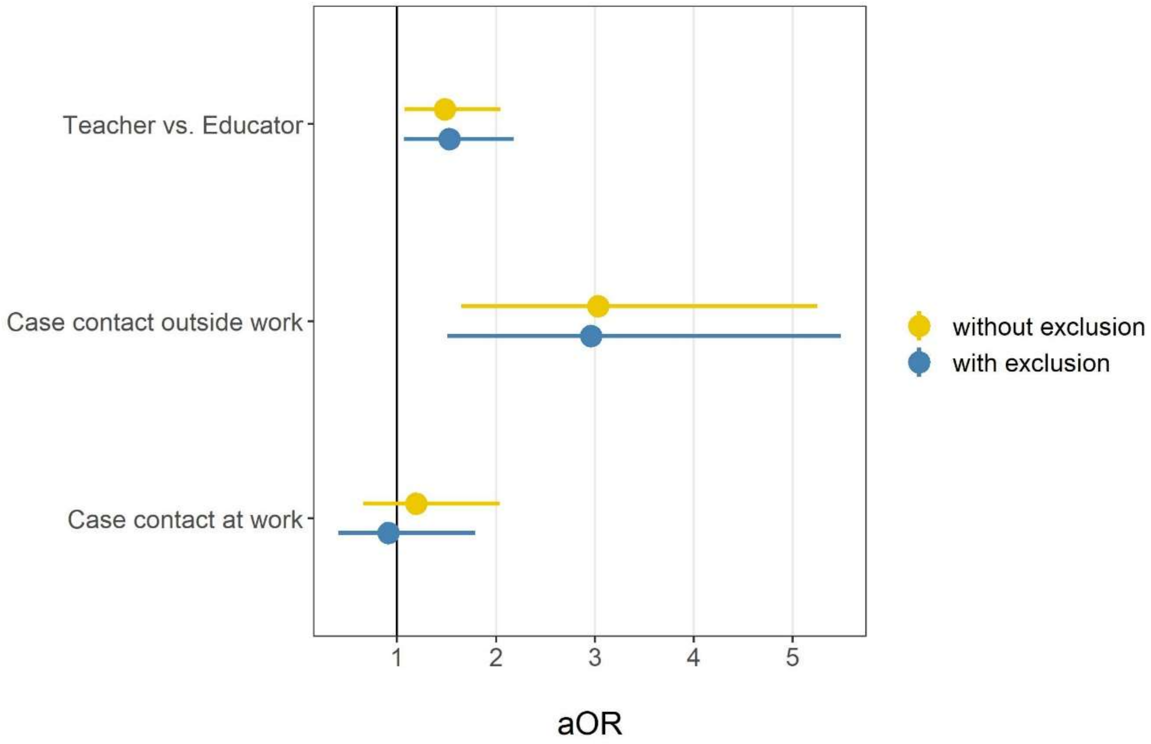
